# Causal Interactions in Brain Networks Predict Pain Levels in Trigeminal Neuralgia

**DOI:** 10.1101/2023.06.02.23290885

**Authors:** Yun Liang, Qing Zhao, John K. Neubert, Mingzhou Ding

## Abstract

Trigeminal neuralgia (TN) is a highly debilitating facial pain condition. Magnetic resonance imaging (MRI) is the main method for generating insights into the central mechanisms of TN pain in humans. Studies have found both structural and functional abnormalities in various brain structures in TN patients as compared with healthy controls. Whereas studies have also examined aberrations in brain networks in TN, no studies have to date investigated causal interactions in these brain networks and related these causal interactions to the levels of TN pain. We recorded fMRI data from 39 TN patients who either rested comfortably in the scanner during the resting state session or tracked their pain levels during the pain tracking session. Applying Granger causality to analyze the data and requiring consistent findings across the two scanning sessions, we found 5 causal interactions, including: (1) Thalamus → dACC, (2) Caudate → Inferior temporal gyrus, (3) Precentral gyrus → Inferior temporal gyrus, (4) Supramarginal gyrus → Inferior temporal gyrus, and (5) Bankssts → Inferior temporal gyrus, that were consistently associated with the levels of pain experienced by the patients. Utilizing these 5 causal interactions as predictor variables and the pain score as the predicted variable in a linear multiple regression model, we found that in both pain tracking and resting state sessions, the model was able to explain ∼36% of the variance in pain levels, and importantly, the model trained on the 5 causal interaction values from one session was able to predict pain levels using the 5 causal interaction values from the other session, thereby cross-validating the models. These results, obtained by applying novel analytical methods to neuroimaging data, provide important insights into the pathophysiology of TN and could inform future studies aimed at developing innovative therapies for treating TN.

## 1 Introduction

Trigeminal neuralgia (TN) is a distressing condition characterized by severe facial pain that is akin to electric shocks and can be triggered by even the slightest touch (Zakrzewska and Linskey, 2014). Its occurrence is estimated to be approximately 4.3 cases per 100,000 individuals annually, with a slightly higher prevalence observed in women (5.9/100,000) compared to men (3.4/100,000) (Obermann, 2010). TN exhibits an age-related pattern, primarily affecting older individuals, while it is relatively rare among those below the age of 40 (Bennetto et al., 2007). Extensive research has delved into understanding the peripheral mechanisms underlying TN pain. Utilizing magnetic resonance imaging (MRI) technology, these investigations have consistently identified a strong correlation between neurovascular contact (NVC) and the symptomatic side in classical TN, specifically in cases where NVC leads to trigeminal nerve displacement or atrophy, distinguishing it from general instances of NVC (Maarbjerg et al., 2015). Notably, NVC at the trigeminal root entry zone (REZ) is particularly likely to manifest as symptomatic TN when accompanied by structural changes in the nerve (Antonini et al., 2014). However, given the relatively high relapse and recurrence for TN following both pharmacological and surgical therapies, a deeper comprehension of the central mechanisms underlying TN pain is necessary to develop more effective therapeutic interventions.

Functionally, researchers have begun to apply functional MRI (fMRI) to understand pain-related brain patterns (Wang et al., 2015; Yan et al., 2019; Zhu et al., 2020), and have consistently observed the activation of a set of brain regions in both chronic and experimental pain (Moisset et al., 2011; Simons et al., 2014). In particular, the importance of the anterior cingulate cortex (ACC) and the thalamus in mediating pain processing is well recognized (Bushnell et al., 2013; Legrain et al., 2011). The thalamus, being a key node in the ascending pain pathway, receives the nociceptive input from the periphery and transmits it broadly to other brain structures, including the ACC and insula (Dostrovsky, 2000). The ACC, especially its dorsal portion (dACC) (Lieberman and Eisenberger, 2015), upon receiving the noxious input from the thalamus, processes its cognitive and motivational significance and contributes to the perception of pain. In this conceptualization, one may expect that the stronger the nociceptive input from the thalamus to the dACC, the higher the perceived pain level. Paralleling the ascending pain pathway, there is also a descending pain inhibitory pathway, which includes both the ACC and the thalamus as key nodes. The role of the descending pain inhibitory pathway is to suppress the transmission of nociceptive input to reduce pain. In the case of TN, the thalamus has been shown to play a significant part in the modulation of nociceptive inputs from the spinal trigeminal nucleus (Ab Aziz and Ahmad, 2006). Based on the assumption that the brain’s response to increased pain involves enhanced inhibition of pain signal transmission (Porreca et al., 2002), it is a reasonable expectation that a higher perceived pain level would correspond to a stronger descending inhibitory influence from the ACC to the thalamus. In addition to the thalamus and ACC, one may postulate that other brain structures such as the caudate nucleus, which is an essential node of the descending inhibitory pathway, play a similar role in regulating the activity along the ascending pain pathway.

Testing these hypotheses requires the assessment of directional connectivity or effective connectivity. Conventional functional connectivity analysis, which is based on the covariation of neural activity across brain regions, is inadequate for this purpose because it does not produce directional information. Granger causality (GC) has been shown to be a major tool to overcome this limitation (Granger, 1969). Granger causality is a statistical concept that is grounded in the notion that if past values of one time series can be used to predict the present values of another time series, then the first time series is said to exert causal influences on the second time series. The roles of the two-time series can be reversed to address the causal influences in the opposite direction. In a GC analysis, autoregressive models are fit to the data recorded from different brain regions, from which causal influences between these regions are then derived (Stramaglia et al., 2016). Multiple pain studies have applied GC to brain imaging data (Seth et al., 2015; Wen et al., 2013b). Huang et al. found that when compared with healthy controls (HCs), patients with migraine without aura (MwoA) showed significantly decreased effective connectivity from right amygdala to right superior temporal gyrus, left superior temporal gyrus and right precentral gyrus (Huang et al., 2021). They further indicated that decreased connectivity in each hemisphere from amygdala to superior temporal gyrus in MwoA patients may represent temporal processing impairment. In a separate study on MwoA, significant differences in Granger causality connections between the right-frontoparietal network and executive control network were observed between patients and healthy controls (Ning et al., 2018). These insights, afforded by the directional information provided by GC, are not possible with the conventional functional connectivity analysis. To date, however, no studies have applied the GC analysis to investigate the causal interactions among different brain regions in TN. In this study, aiming for a deeper understanding of the central mechanisms of TN pain, we examined brain network dynamics underlying TN pain with particular focus being placed on how causal interactions in brain networks give rise to TN pain generation and perception.

Functional magnetic resonance imaging (fMRI) data were recorded from TN patients who (1) rated their spontaneous pain levels in the pain tracking session and (2) rested comfortably in the resting state session. Three GC analyses were performed. In the first analysis, dACC and the thalamus were chosen as the regions of interest (ROIs). Their causal interactions were evaluated in both pain tracking and resting state sessions and correlated with the pain levels. In the second analysis, the list of ROIs was expanded to include the caudate, anterior insula, and primary somatosensory cortex (S1), which, along with dACC and thalamus, are often collectively called the pain matrix. Causal interactions among these ROIs were evaluated and correlated with pain levels in both sessions. In the third analysis, we further expanded the list of ROIs to include 23 regions that have been identified in a recent study as being specific to TN pain (Liang et al., 2022). In all three analyses consistent findings between the resting and the pain tracking sessions were sought to enhance the statistical rigor of the analysis.

## 2 Materials and Methods

This study is a reanalysis of previously published data (Liang et al., 2022). The questions addressed here are, however, entirely different from the previously published work.

### 2.1 Participants

The study was approved by the WCG Institutional Review Board (IRB). In total, 55 patients gave written informed consent and participated in the study (69% female, mean age ± standard deviation (SD)=53.9±14.9). Sixteen patients were rejected due to a combination of the following reasons: (1) not meeting diagnosis criteria (*n=1*), (2) not completing the whole experiment (*n=11*), (3) technical difficulties during fMRI recording (*n=2*), and (4) excessive movements inside the scanner (*n=2*). The data from the remaining 39 patients, who were all diagnosed as having TN1, were analyzed and reported here.

### 2.2 Experimental Paradigm

Prior to neural data recording, the patients indicated on two visual analog scales (VAS): their daily experienced pain level (henceforth referred to as usual pain) and current pain level (henceforth referred to as pain intensity), with 0 indicating no pain and 100 the most excruciating pain. Following that, the patients underwent functional, structural, and diffusion magnetic resonance imaging. As shown in **Figure 1**, there are two types of functional scans: resting state and pain tracking. During the resting state scan (7 minutes), the patients were instructed to fixate the cross at the center of the monitor screen, stay still, and not to think any systematic thought. During the pain tracking scan (10-20 minutes), fMRI data were acquired while the patients rated their pain levels continuously using a tracking ball. The tracking ball controlled the movement of a cursor along a straight line with 0 and 100 indicated at the two ends of the line on the monitor. It is worth noting that for 7 of the 39 patients, the pain tracking session was 10 minutes in duration (Liang et al., 2022).

**Figure 1.**
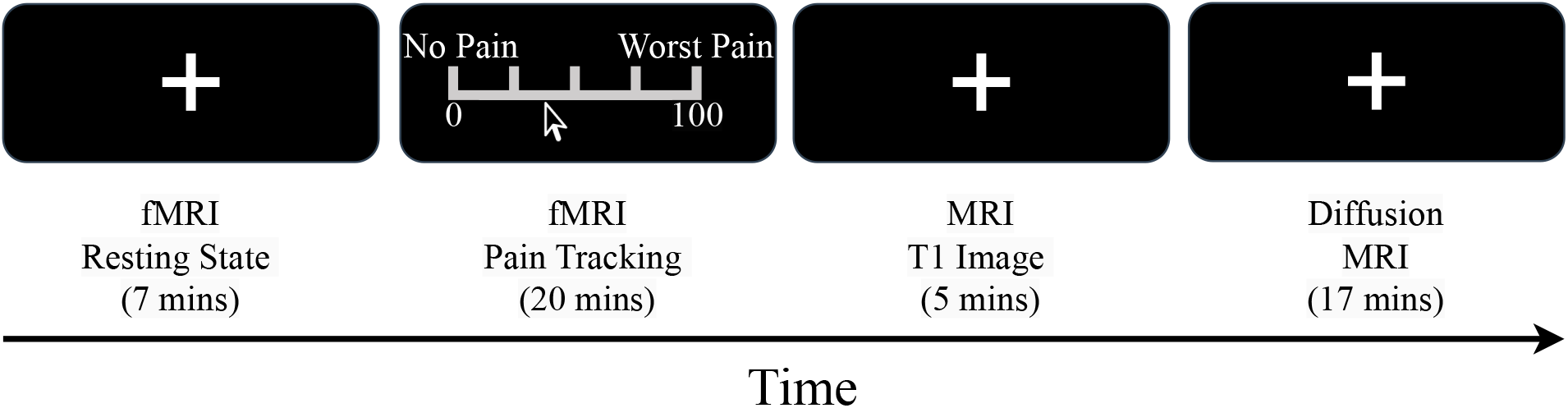
Study design. The experiment was divided into four sessions: resting-state, pain tracking, T1 imaging, and diffusion MRI.

### 2.3 Functional MRI Data Acquisition and Preprocessing

Functional MRI images were collected on a 3 T Philips Achieva scanner (Philips Medical Systems, the Netherlands) equipped with a 32-channel head coil. The echo-planar imaging (EPI) sequence parameters were as follows: repetition time (TR), 1.98 sec; echo time, 30 msec; flip angle, 80; field of view, 224 mm; slice number, 36; voxel size, 3.5×3.5×3.5 mm; matrix size, 64×64. In addition to the functional scans, a high-resolution anatomical T1-weighted MRI image was also acquired for each subject using the following parameters: field of view (FOV), 240×240 mm; time of repetition (TR), 8.0566 ms; time of echo (TE), 3.686 ms; resolution, 1×1 mm; flip angle, 80 degrees. EEG data were concurrently collected with fMRI but not analyzed here. Likewise, diffusion MRI was also recorded but not analyzed here.

Statistical Parametric Mapping (SPM) was used to preprocess the fMRI data. The preprocessing steps include slice timing correction, realignment, co-register, spatial normalization, and spatial smoothing. Data segments with large head movements were removed from 5 patients. For the fMRI data that were subjected to further analysis, head motions whose magnitude were within acceptable range, were regressed out, and a high-pass filter with a cutoff frequency set at 0.01 Hz was applied to attenuate lower frequency noise.

### 2.4 Definition of ROIs

#### Analysis 1: dACC and thalamus

In the first analysis, we examined how causal interactions between dACC and the thalamus are related to TN pain experienced by the patient. The dACC ROI was based on the Lausanne Atlas (Hagmann et al., 2008). The Oxford Thalamic Connectivity Probability Atlas was used to extract the thalamus ROI (Behrens et al., 2003). Specifically, the ventral posterior region of the thalamus, which is thought to be related to the human ventral posterior nucleus (VP), was chosen as our thalamic ROI (Behrens et al., 2003). This region has been used as the somatosensory thalamus in previous studies (Cunningham et al., 2017).

#### Analysis 2: The pain matrix

Numerous neuroimaging studies have consistently identified a set of brain regions, including dACC, thalamus, caudate, S1 and anterior insula as playing an important role in both chronic and experimental pain (Cacioppo et al., 2013; Wilcox et al., 2015). These regions, collectively referred to as the pain matrix, constitute the ROIs in our second analysis. We examined the pairwise causal interactions among the five pain matrix regions and their relationship with TN pain experienced by the patient. The caudate ROI, anterior insula ROI, and SI_face were extracted from the Lausanne atlas (Hagmann et al., 2008).

#### Analysis 3: 23 TN pain related regions

In our previous study, applying correlation analysis, convolutional neural network (CNN) analysis, and graph convolutional neural network (GCNN) analysis (Liang et al., 2022), and requiring consistent results across multiple methods, we have identified 23 brain regions that are related to the generation and perception of TN pain; see **Table 1**. In the third analysis, we examined the causal interactions in this TN pain network and how these causal interactions related to TN pain experienced by the patient.

**Table 1:**
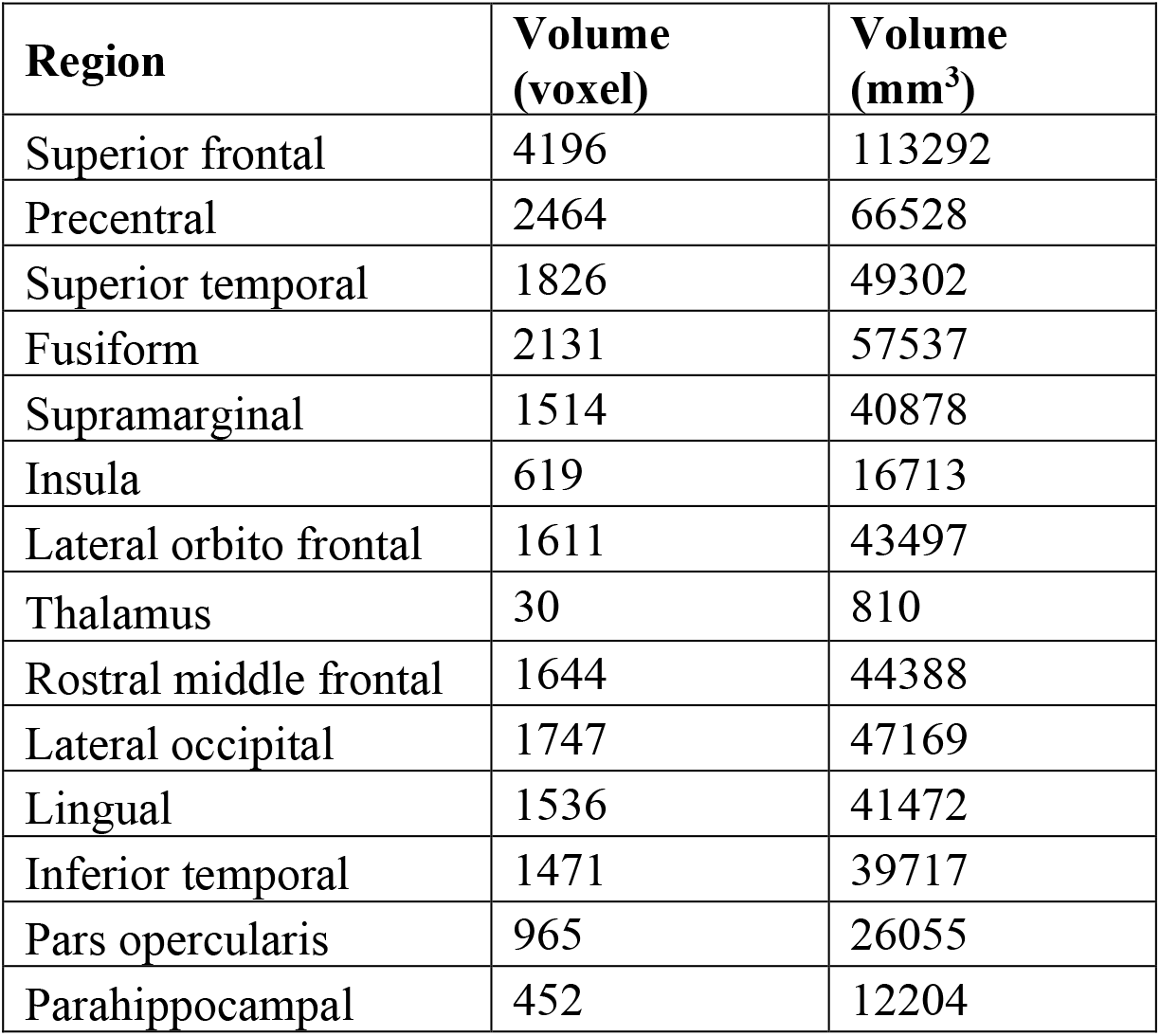
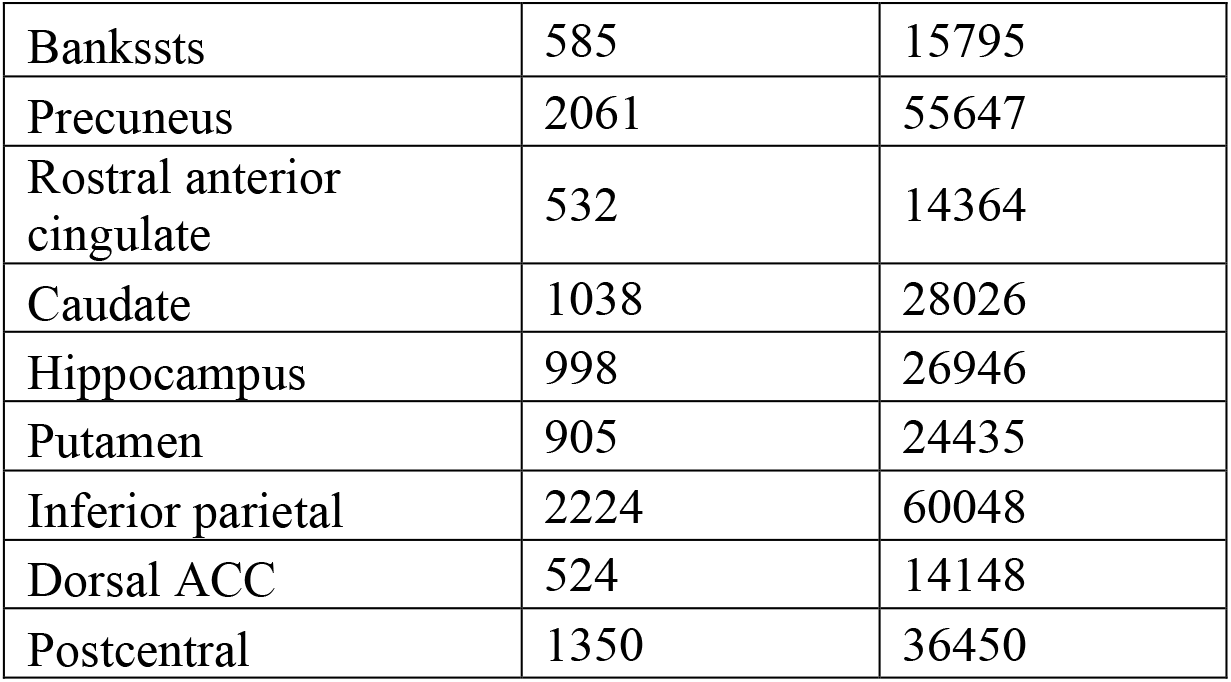
Regions in the TN pain network identified by correlation and AI-inspired analyses in our previous study (Liang et al., 2022). The volume size for each ROI was listed in the table.

| Region | Volume (voxel) | Volume (mm<sup>3</sup>) |
| --- | --- | --- |
| Superior frontal | 4196 | 113292 |
| Precentral | 2464 | 66528 |
| Superior temporal | 1826 | 49302 |
| Fusiform | 2131 | 57537 |
| Supramarginal | 1514 | 40878 |
| Insula | 619 | 16713 |
| Lateral orbito frontal | 1611 | 43497 |
| Thalamus | 30 | 810 |
| Rostral middle frontal | 1644 | 44388 |
| Lateral occipital | 1747 | 47169 |
| Lingual | 1536 | 41472 |
| Inferior temporal | 1471 | 39717 |
| Pars opercularis | 965 | 26055 |
| Parahippocampal | 452 | 12204 |
| Bankssts | 585 | 15795 |
| Precuneus | 2061 | 55647 |
| Rostral anterior cingulate | 532 | 14364 |
| Caudate | 1038 | 28026 |
| Hippocampus | 998 | 26946 |
| Putamen | 905 | 24435 |
| Inferior parietal | 2224 | 60048 |
| Dorsal ACC | 524 | 14148 |
| Postcentral | 1350 | 36450 |

### 2.5 GC analyses

GC is a statistical method for assessing the causal relations between two simultaneously recorded time series (e.g., the BOLD time courses from two ROIs) (Ding et al., 2006; Granger, 1969). Consider one of the time series. Fit an autoregressive model on the data and assess the variance of the prediction error. Then fit an autoregressive model on both time series data. If the variance of the prediction error of the first time series is reduced by using the past measurements of the second time series, then we say that there is a causal influence from the second time series on the first time series. The role of the two-time series can be reversed to evaluate the causal influence from the first series on the second one. For this study, consider two ROIs (e.g., dACC and thalamus). First, the pre-processed Blood Oxygenation Level Dependent (BOLD) time series from all voxels within a ROI was averaged to yield one time series. Second, the temporal mean was removed from the time series to meet the zero-mean requirement assumed by the autoregressive (AR) model used to estimate GC (Ding et al., 2000; Ding et al., 2006). Third, because of the assumption of stationary times series in Granger causality analysis, we tested the stationarity of the BOLD time series by the augmented Dickey-Fuller test. All the time series analyzed here meet the stationarity condition. Fourth, bivariate AR models were fitted for the BOLD time series from a pair of ROIs and the GC values from the two directions (ROI 1→ROI 2 and ROI 2→ROI 1) were derived. A mode order of 7 was chosen based on our recently proposed criterion where the difference between the spectral estimates of the BOLD time series from the AR model and that from the Fourier method was minimized (Rajan et al., 2019; Trongnetrpunya et al., 2016; Wang et al., 2016a).

## 3 Results

In this work, causal connectivity analyses were applied to fMRI data recorded from TN patients in two recording sessions: pain tracking session and resting state session. The goal was to examine whether causal interactions in brain networks are associated with pain levels experienced by the patients. Consistent findings across the two recording sessions were emphasized to enhance statistical robustness.

### 3.1 **Pain rating analysis**

Three pain ratings were obtained from each patient: the usual pain (the daily experienced pain level), the pain intensity (the pain level experienced right before the experiment), and the average pain rating (the average level of pain experienced by the patient during the pain tracking session). Significant differences were observed between usual pain and average pain ratings (p<0.0001), as well as between usual pain and pain intensity (p<0.0001), but not between pain intensity and average pain ratings (p>0.05), as depicted in **Figure 2(A)**. These results are expected given that (1) the pain levels can fluctuate significantly over an extended period of time and (2) the pain intensity right before the experiment and the average pain ratings during the experiment were collected close together in time. We also found significant correlations between usual pain and pain intensity (R=0.34, p=0.03) as well as between usual pain and average pain ratings (R=0.38, p=0.02). To extract a single pain score from the three pain ratings, a principal component analysis (PCA) was carried out, and the first PCA component was found to explain 61% of the variance (**Figure 2(B))**. The score on the first PCA component, referred to as the pain score henceforth, was used to represent the pain level of each patient in subsequent sections.

**Figure 2.**
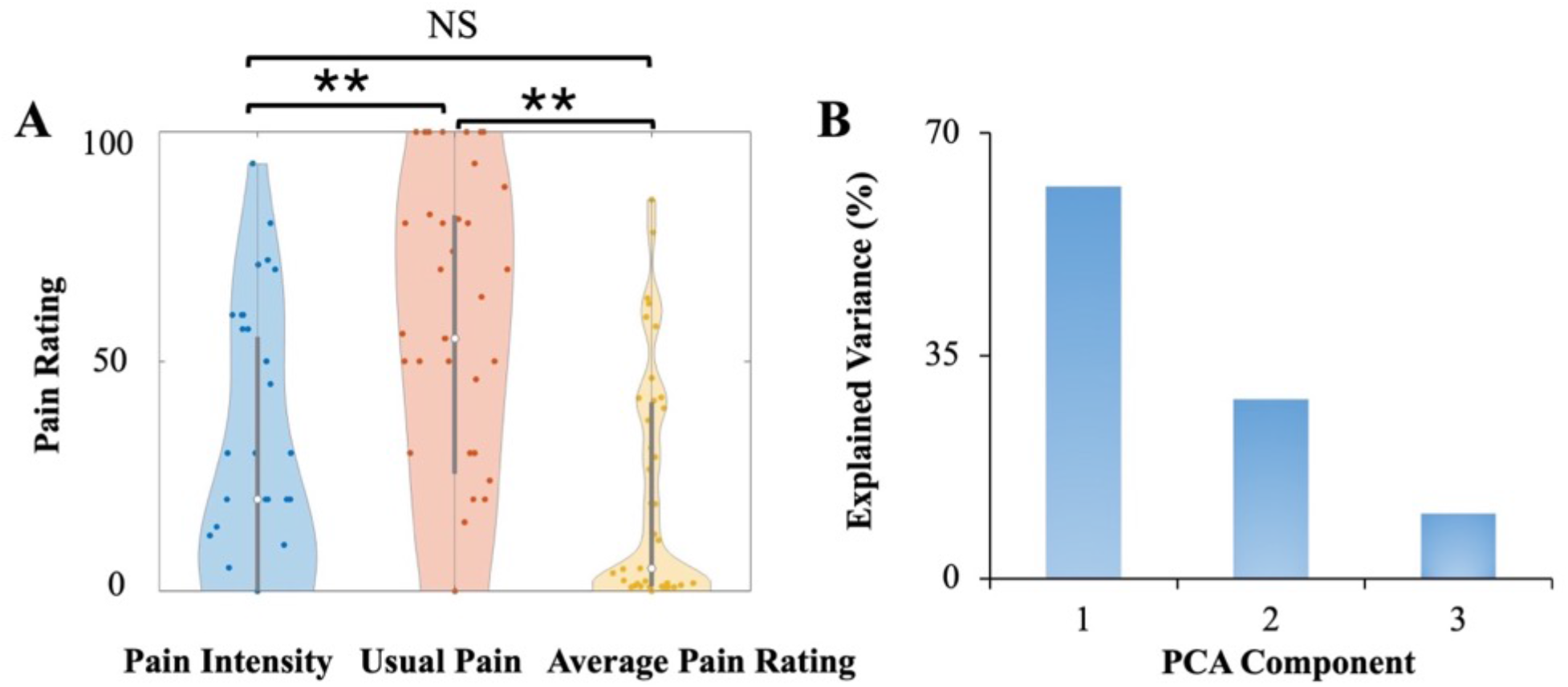
Pain rating analysis. **(A)** The three pain ratings included in the study: Pain Intensity, Usual Pain, and Average Pain Rating. Significant differences were observed between usual pain and average pain ratings (p<0.0001), as well as between usual pain and pain intensity (p<0.0001), but no significant difference was found between pain intensity and average pain ratings (p>0.05). **(B)** The variance explained by each of the three PCA components. The first PCA component explained 61% of the total variance. ** p<0.0001.

### 3.2 Analysis 1: Causal interactions between thalamus and dACC and TN pain

The two ROIs considered in this analysis were illustrated in **Figure 3(A)**. Scatter plots in **Figure 3(B)** show a significant association between Thalamus **→** dACC causal connectivity and pain score in two sessions (R=0.45, p=0.004; R=0.34, p=0.032). For the correlation between dACC **→** Thalamus and pain score, however, we noted that the results were severely influenced by one outlier in both sessions, and thus excluded this direction from further consideration. **Figure 3(C)** and **3(D)** summarize these results in a matrix form where the causal influence is from the Source to the Target (Source **→** Target). These correlation results suggest that stronger nociceptive input from the thalamus to the dACC is associated with higher perceived pain levels, supporting our hypothesis. We schematically represent this relationship in **Figures 3(E)** and 3**(F)**.

**Figure 3.**
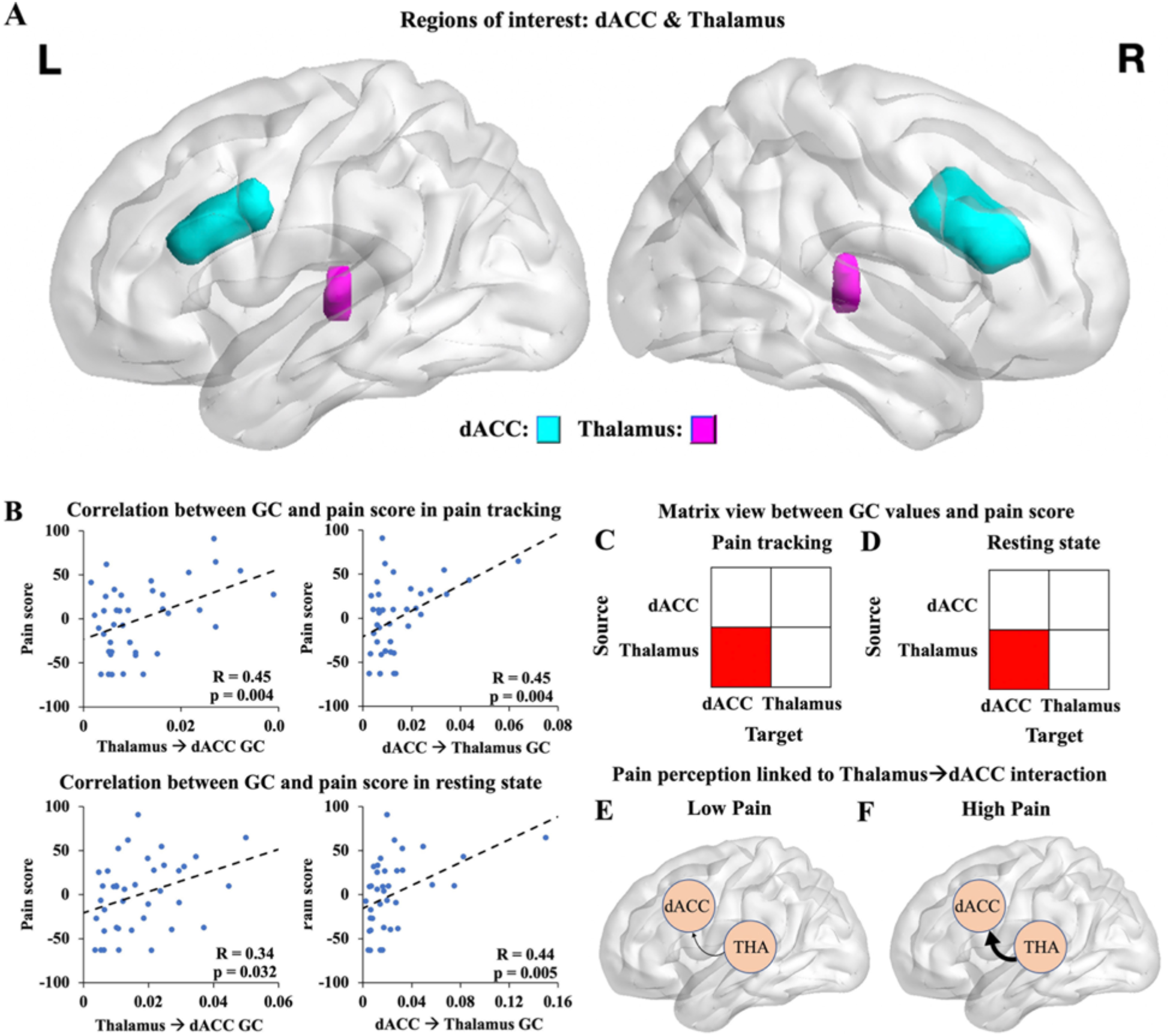
GC between thalamus and dACC and pain score. **(A)** The ROIs: dACC (blue); thalamus (pink). **(B)** Correlation between Thalamus→dACC and pain score and between dACC→Thalamus and pain score in pain tracking and resting state sessions. It can be seen that Thalamus**→**dACC was consistently correlated with pain score in both sessions. The correlation between dACC**→**Thalamus and pain score appeared to be driven by an outlier in both sessions and was removed from further consideration. **(C)-(D)** The correlation results from the two sessions summarized in a matrix form. A cell was colored red if Source**→**Target is significantly correlated with pain score at p<0.05; otherwise, it was left blank. **(E)-(F)** Schematic to illustrate that the stronger the nociceptive input from the thalamus to the dACC, the higher the perceived pain level. dACC: dorsal anterior cingulate cortex.

### 3.3 Analysis 2: Causal interactions in the pain matrix and TN pain

In this analysis, we expanded beyond the thalamus and dACC by including S1 (face), caudate, and anterior insula, which are other major areas in the pain matrix, as additional ROIs; see **Figure 4(A)**. These areas have all been previously implicated in pain processing (Fomberstein et al., 2013; Moisset et al., 2011). We computed pairwise GC among all pain matrix regions during the pain tracking and resting sessions and correlated the GC results with the pain score. In the pain tracking session, in addition to Thalamus **→** dACC studied above, Caudate **→** S1 (R=0.34, p=0.035), and Thalamus **→** S1 (R=0.40, p=0.013) showed significant correlations with the pain score, as depicted in **Figure 4(B)**. In the resting state, in addition to Thalamus **→** dACC studied above, GC from Insula **→** Thalamus (R=0.37, p=0.020), S1 **→** Insula (R=0.40, p=0.011), and S1 **→** Thalamus (R=0.33, p=0.039) showed significant correlations with the pain score, as depicted in **Figure 4(C)**. However, it remains that Thalamus **→** dACC is the only causal interaction that showed consistent association with the pain score across the two recording sessions.

**Figure 4.**
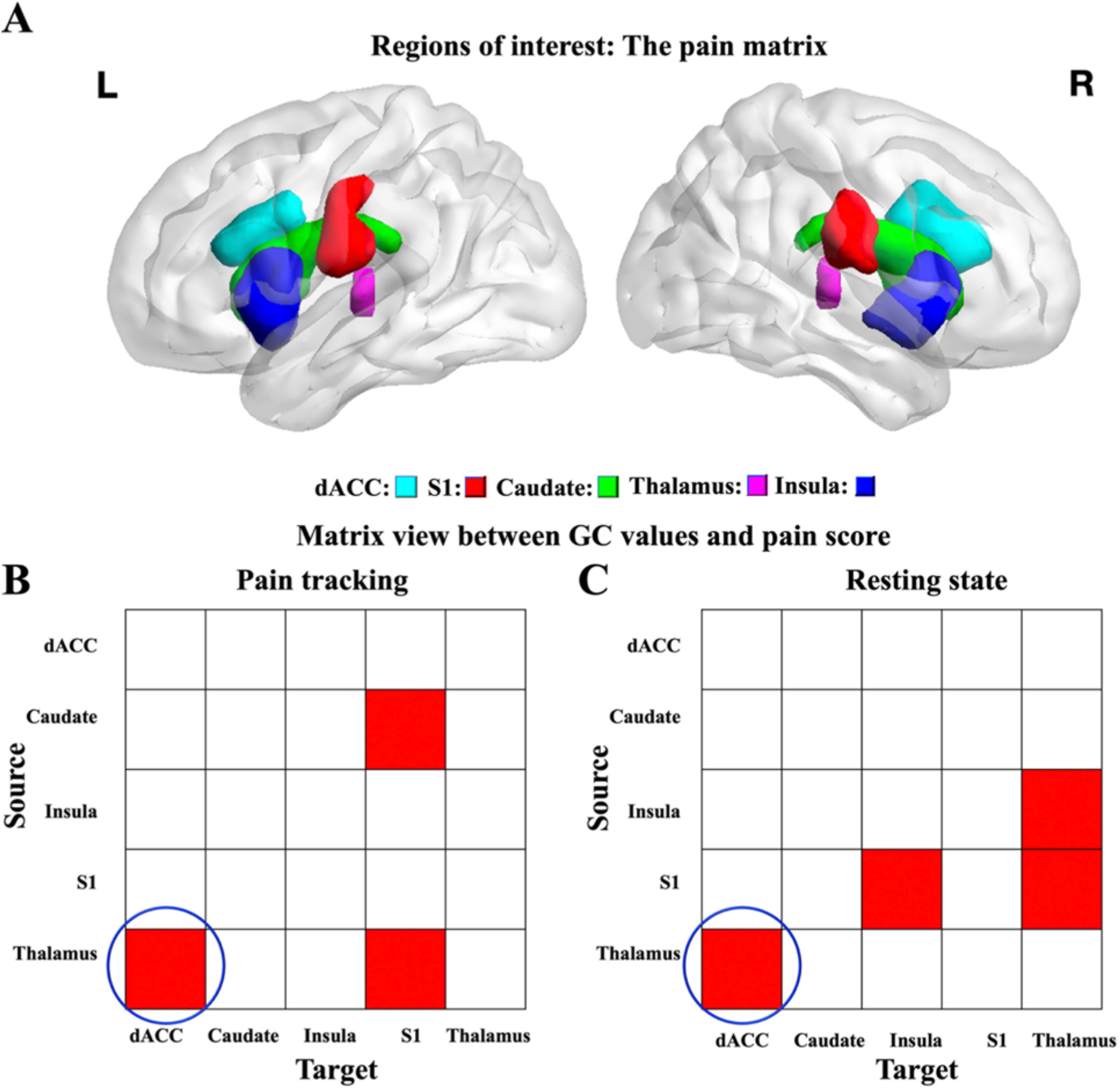
GC in the pain matrix and pain score. **(A)** The ROIs: dACC (blue); S1 face area (red); Caudate (green); Thalamus (pink); Insula (navy blue). **(B)-(C)** Matrix view of the relationship between GC values and the pain score for all ROI pairs. Red cells indicate significant correlations between Source**→**Target and pain score at p<0.05; otherwise, it was left blank (white). Only the causal interaction from Thalamus**→**dACC is significantly correlated with the pain score consistently across the two sessions (highlighted by blue circles). dACC: dorsal anterior cingulate cortex; S1: primary somatosensory cortex (face).

### 3.4 Analysis 3: GC in TN pain network and TN pain

In a recent study, employing a variety of methods, we found 23 brain regions whose activities are related to the generation and perception of TN pain (Liang et al., 2022); see Table 1 and **Figure 5(A)**. These 23 regions were collectedly referred to as the TN pain network here. We computed the pairwise GC in the TN pain network in the pain tracking and resting state sessions and related that to the pain score. As shown in **Figure 5(B) and 5(C)**, we found that besides the causal connectivity from Thalamus **→** dACC, additional causal influences, including Caudate **→** Inferior temporal gyrus, Precentral gyrus **→** Inferior temporal gyrus, Supramarginal gyrus **→** Inferior temporal gyrus and Bankssts **→** Inferior temporal gyrus, were significantly and consistently correlated with the pain score across the two sessions (matrix elements consistently marked red in the two recording sessions highlighted by blue circles).

**Figure 5.**
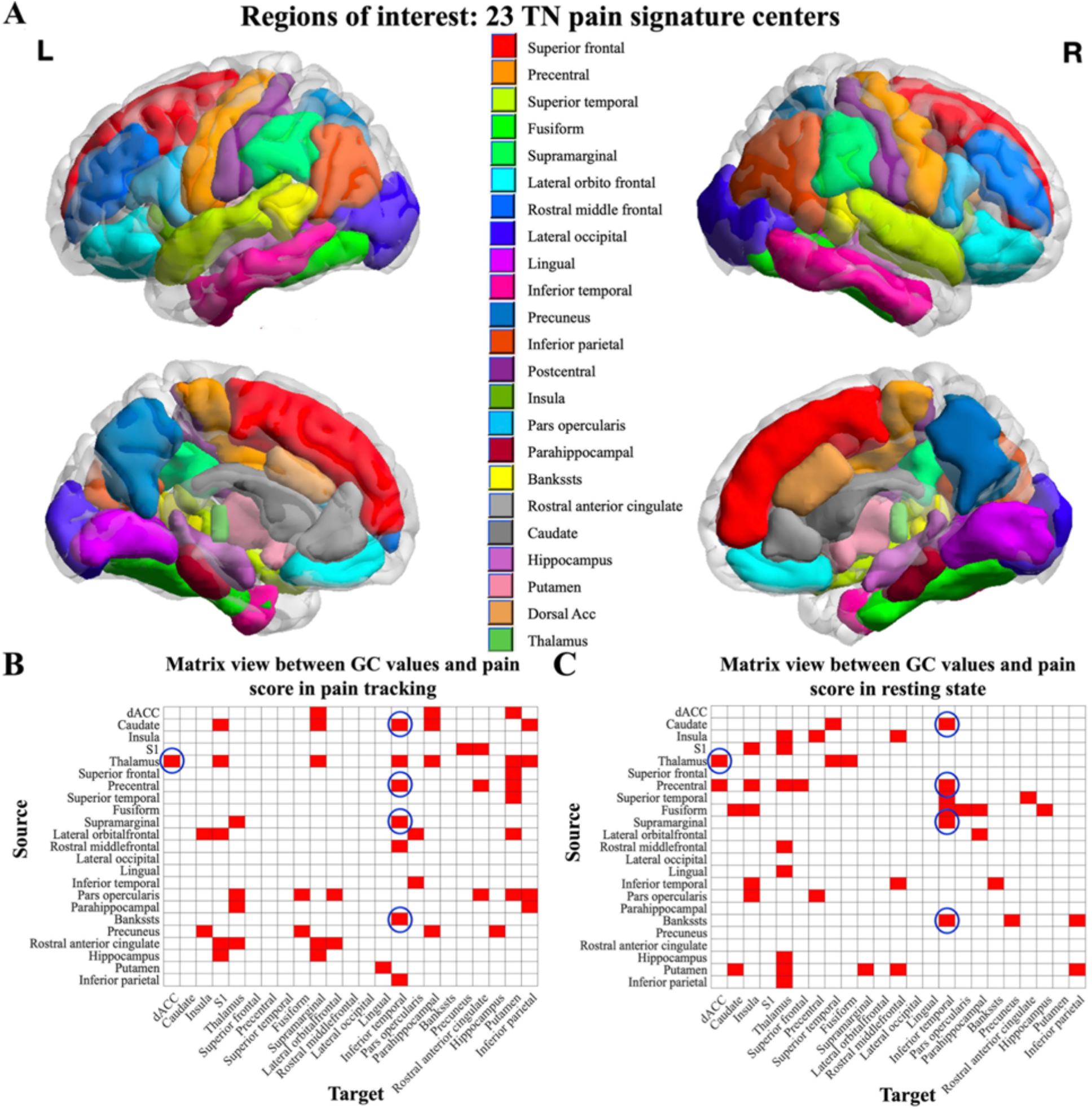
GC in the TN pain network and the pain score. **(A)** 23 TN pain signature center ROIs visualized on the brain. Matrix view of the relationship between GC values and the pain score for all 23 ROI pairs in **(B)** pain tracking and **(C)** resting session with the pain score. A red colored cell denotes a significant correlation (p<0.05); non-significant correlations are denoted in white. The five consistent directions that showed significant GC-pain score correlations in both sessions are highlighted with blue circles. dACC: dorsal anterior cingulate cortex; S1: primary somatosensory cortex (face).

### 3.5 Causal interactions in TN pain network and pain score: Multiple regression analysis

The five causal interactions within the TN pain network that were consistently related to the pain score were shown in **Figure 6(A)**. To examine the collective relation of these causal interactions with the pain score, we constructed a multiple regression model using the pain score as the predicted variable and the 5 causal interactions as the predictor variables, as shown in **Figure 6(B)**. In **Figure 6(C)**, a strong correlation was observed between the predicted pain score and the actual pain score in the pain tracking session (R=0.60, p<0.0001) and in the resting state session (R=0.60, p<0.0001). To cross validate the models, the multiple regression model obtained from one session was tested on the data from the other session. As shown in **Figure 6(D)**, applying the model trained on the pain tracking session to the data in the resting session yielded a significant correlation between the predicted and actual pain scores (R=0.53, p=0.0006), and applying the model trained on the resting session to the data in the pain tracking session resulted in a significant correlation (R=0.54, p=0.0004). These cross-validation results underscore the robustness and generalizability of our predictive model.

**Figure 6.**
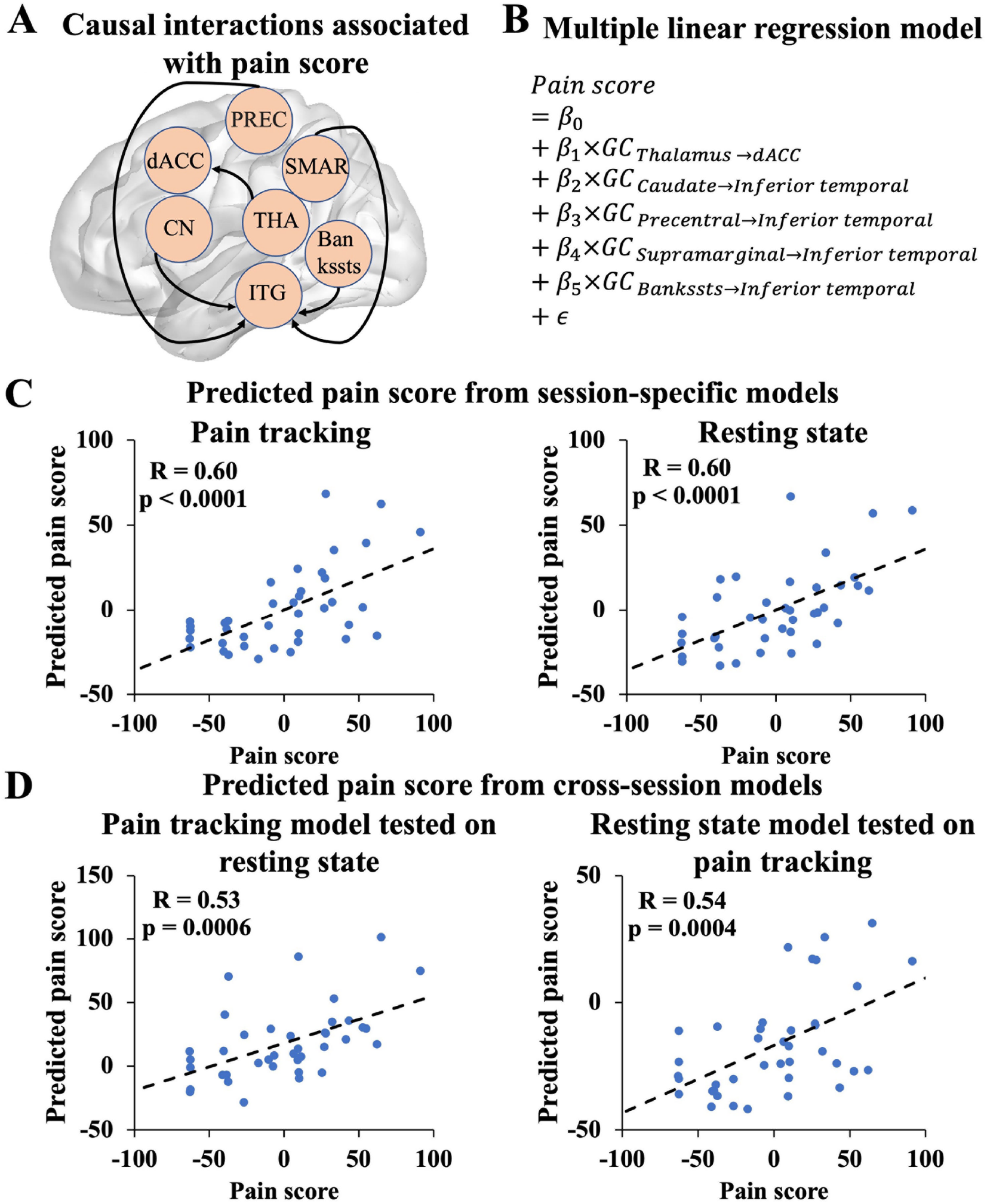
Multiple linear regression models relating causal interactions in brain networks to TN pain. **(A)** Five causal interactions consistently associated with TN pain. dACC: Dorsal anterior cingulate cortex; PREC: Precentral gyrus; SMAR: Supramarginal gyrus; CN: Caudate nucleus; THA: Thalamus; ITG: Inferior temporal gyrus. **(B)** The multiple regression model with the pain score as the predicted variable and the five causal interactions as the predictor variables. **(C)** The pain score vs. predicted pain score in pain tracking session and resting state session. **(D)** Cross validation of the multiple regression model: the model fitted on data from one session was tested on data from the other session. GC: Granger causality.

## 4. Discussions

fMRI data from TN patients were recorded in two scanning sessions: pain tracking and resting state. We examined whether the level of pain experienced by the patient can be linked to causal interactions in brain networks. Starting with dACC and the thalamus, where clear hypotheses can be proposed based on these two brain areas’ known functions in pain processing, we showed that Thalamus **→** dACC is consistently associated with pain levels across the two scanning sessions. Expanding beyond dACC and thalamus, we included three additional areas from the so-called pain matrix: caudate, S1 (face), and anterior insula. Among the pairwise causal interactions among the five ROIs in the pain matrix, Thalamus **→** dACC remained to be the only causal interaction that is consistently correlated with the pain score across the two scanning sessions. Further expanding the investigation to consider 23 brain regions that have been identified in our recent study as related to TN pain (Liang et al., 2022), we discovered a total of 5 causal interactions that were significantly and consistently correlated with the pain score: Thalamus **→** dACC, Caudate **→** Inferior temporal gyrus, Precentral gyrus **→** Inferior temporal gyrus, Supramarginal gyrus **→** Inferior temporal gyrus and Bankssts **→** Inferior temporal gyrus. Utilizing these 5 causal interactions as predictor variables and the pain score as the predicted variable in a linear multiple regression model, we found that in both pain tracking and resting state sessions, the model was able to predict the pain levels and explain ∼36% of variance. More importantly, the model trained on the 5 causal interaction values from one session was able to predict pain levels using the 5 causal interaction values from the other session, thereby cross-validating the models. To the best of our knowledge, this is the first study that utilized Granger causality analysis to relate causal interactions in brain networks to TN pain.

Previous studies on pain processing have identified two ascending pain-supporting pathways and one pain-inhibitory descending pathway (De Ridder and Vanneste, 2016). The lateral ascending pain pathway is responsible for processing the discriminatory components of pain, while the medial ascending pain pathway is responsible for the motivational, affective, and attentional components of pain (De Ridder and Vanneste, 2016). These pathways operate in parallel and can be independently modified (Frot et al., 2008). The thalamus, as part of the ascending pain pathways, receives nociceptive input from the periphery (Ab Aziz and Ahmad, 2006) and processes the nociceptive information before transmitting it to the cerebral cortex. This central importance of the thalamus in pain processing is the reason for us to choose the thalamus as one of the key ROIs in the present analysis. The dACC, another key ROI in the present study, is also known to play a crucial role in pain perception and modulation (Eisenberger, 2012). Activation in the dACC in response to painful stimuli is thought to reflect the detection of nociceptive input from ascending pain pathways, particularly from the thalamus (Yin et al., 2020). Moreover, the magnitude of pain-evoked activation in the dACC is deemed crucial in determining an individual’s emotional and behavioral reactions to pain (Rainville et al., 1997). A study goes as far as to suggest that the dACC’s main function is pain processing (Lieberman and Eisenberger, 2015) despite its known involvement in a broader array of executive, cognitive and sensory functions (Davis et al., 1997). dACC’s top-down regulatory functions have also been considered in recent studies including its suppression of the brain default mode activity during attention tasks (Rajan et al., 2019; Wen et al., 2013a). In our study, we observed that the causal influence from the thalamus to dACC, denoted as Thalamus **→** dACC, was positively associated with the level of TN pain, further confirming a role of these two regions in TN pain generation and perception. The causal influence in the reverse direction, dACC **→** Thalamus, was not found to be predictive of pain levels, suggesting that dACC may not play a role in the downregulation of thalamic activity in TN pain. These results, highlighting the causal interplay between two critical structures in the processing of TN pain, may have implications for the identification of potential targets for novel therapies.

The primary somatosensory cortex (S1), insula, and caudate nucleus, along with the thalamus and ACC, have been collectively referred to as the pain matrix (DeCharms et al., 2005). Lesions within S1 result in deficits in detecting sensory features of pain in humans (ECHOLS and Colclough, 1947; Greenspan et al., 1999), and neural activity in S1 closely correlates with the temporal and intensity aspects of pain sensations (Coghill et al., 1999; Porro et al., 1998). Specific to TN pain, a recent fMRI study found that several structures, including S1 and insula, were activated in patients with classical TN without evoked pain (Moisset et al., 2011). The caudate nucleus, a subcortical structure, is a major component of the descending pain-inhibition system and has been implicated in modulating chronic pain perception (Li et al., 2017b; Wunderlich et al., 2011; Yang et al., 2010). Studies have indicated that activation of the caudate nucleus occurs during control and suppression of painful stimuli (Freund et al., 2009; Freund et al., 2007). Researchers have also suggested that caudate activation is a distinguishing feature in the early phase of the tonic painful electrostimulation during the task to suppress the feeling of pain (Wunderlich et al., 2011). Regarding TN, a recent voxel-based morphometry (VBM) study detected reductions in gray matter volume in the caudate nucleus in TN patients, further implicating its involvement in TN pathology (Li et al., 2017a). The insula’s role in pain processing is well recognized. It has been implicated in both pain perception and pain modulation (Ichesco et al., 2016). Prior research has established that the medial pain system mediates the more affective-motivational aspects of the experience of pain, and this circuitry mainly relays information through the thalamus to the anterior insula and ACC, integrating interoceptive input with its emotional salience (Taylor et al., 2009). A recent combined structural and functional connectivity analysis of insular subdivisions supported that the anterior insular is associated with cognitive-affective dimension of pain (Wiech et al., 2014). In the case of TN pain, Borsook et al. (Borsook et al., 2007) reported increased insular activation during both evoked and spontaneous tics compared to healthy controls. Given the foregoing, one may expect additional causal interactions besides Thalamus **→** dACC that should be linked to TN pain. In our second analysis, Caudate **→** S1 was indeed found to be significantly correlated with the pain score in the pain tracking session; however, such relation was not found in the resting state session. Similarly, Insula **→** Thalamus was found to be significantly correlated with the pain score in the resting state session, but not in the pain tracking session. Thus, among the pain matrix ROIs, Thalamus **→** dACC remained to be the causal interactions that was significantly correlated with the pain score consistently across the two scanning sessions.

Our third analysis included 23 brain regions which we have identified in a recent study as being related to TN pain (Liang et al., 2022). In addition to Thalamus **→** dACC, four additional causal interactions were found to be consistently correlated with the pain score across the two scanning sessions, including Caudate **→** Inferior temporal gyrus, Precentral gyrus **→** Inferior temporal gyrus, Supramarginal gyrus **→** Inferior temporal gyrus, and Bankssts **→** Inferior temporal gyrus. Of note, these four causal interactions, stemming from four different source regions, share the same target region – inferior temporal gyrus. The inferior temporal gyrus (ITG) has been established as a node of the default mode network (DMN), a resting-state network that is critical for self-cognition, emotional processing, and memory (Chen et al., 2019; Zhang et al., 2010). Given its involvement in these functions, the DMN has been proposed to associate environmental stimuli (Raichle and Gusnard, 2005), including pain (Otti et al., 2010), with personal experience and memory (Kim, 2010; Sestieri et al., 2011). In the context of chronic pain, alterations in the function of DMN have been observed. Specifically, increased intrinsic DMN activity and enhanced connectivity between the DMN and insula have been associated with heightened clinical pain in individuals suffering from chronic pain conditions such as low back pain, fibromyalgia, and migraine (Coppola et al., 2018; Loggia et al., 2013). Furthermore, resting-state connectivity between the DMN and the anterior insula has been shown to encode the severity of clinical pain, demonstrate sensitivity to fluctuations in pain within individuals, and predict persistent clinical pain (Kim et al., 2019; Loggia et al., 2013). As a critical node of DMN, recent research has also suggested that abnormal neuronal activity in the ITG may also play a role in the mechanism of TN pain. For instance, Xiang et al. reported that patients with chronic TN had elevated regional homogeneity (ReHo) values in the right ITG, among other brain regions, indicating that chronic TN may affect the function of the ITG (Xiang et al., 2019). This finding is consistent with the report by Wang et al. (Wang et al., 2015) suggesting that chronic TN may impact the function of the ITG. In our study, we identified ITG as a crucial target ROI for causal influences from other brain structures such as the caudate, supporting the notion that the ITG may serve as the gateway for pain-related signals to reach the DMN to heighten the focus on pain-related experiences and rumination.

Regarding analytical methodology, past fMRI functional connectivity studies on TN pain have mainly relied on undirected correlation measurements (i.e., conventional functional connectivity) (Tian et al., 2016; Tsai et al., 2018; Tu et al., 2019; Wakaizumi et al., 2019). For instance, one study on TN utilized seed-based correlation analysis with the entire brain using seed regions derived from VBM analysis (Tsai et al., 2018), and demonstrated that the salient network, including the anterior insula and dACC, plays a role in integrating information related to impending stimulation into perceptual decision-making in the context of pain (Tsai et al., 2018). In another study, distinct abnormalities in large-scale brain networks, such as DMN, sensory-motor network, and salience network, were found to be relevant to facial pain in patients with chronic TN, and no significant abnormalities were observed in other networks such as the dorsal attention network, suggesting that the disorder selectively impacts major brain networks (Xu et al., 2022). Despite the significant insights generated by these analyses, using undirected functional connectivity, we are not able to further dissect the specific role of each brain region in a given functional interaction, limiting our ability to test hypotheses that posit different roles for information following in different directions. Consider two ROIs A and B. When A and B functionally interact (i.e., their activities are correlated), the interaction can have three components: A **→** B, B **→** A, and common input that influences both A and B. Granger causality is a method that is able to achieve functional decomposition of the interaction into its distinct components, which is crucial for testing hypotheses that specify different roles of ROIs involved in given functional interactions, and has thus been used extensively in basic neuroscience (Bollimunta et al., 2011; Wen et al., 2012). GC’s application in clinical neuroscience is also increasing with diseases investigated with the method ranging from stroke (Zhao et al., 2016) to Alzheimer’s disease (Chen et al., 2016) to migraine (Ning et al., 2018; Seth et al., 2015; Wang et al., 2016b; Wen et al., 2013b). For our study, when considering the dACC and thalamus and applying an undirected correlation, the functional connectivity between dACC and thalamus is not significantly and consistently correlated with the pain score across the two scanning sessions. Based on the statistical robustness criteria mentioned earlier, from a brain network standpoint, these two ROIs will not be considered important in TN pain generation and perception. However, after we decomposed the interaction into its directed components, we are able to identify Thalamus **→** dACC as being the component of dACC-thalamus interaction that robustly drives the level of the experienced pain level, thereby yielding mechanistic insights not possible with conventional functional connectivity analysis.

In conclusion, in this study, we applied Granger Causality to fMRI data to investigate how causal interactions in brain networks are related to pain levels experienced by TN patients. Five such causal interactions were robustly identified, based on which, a multiple regression model was estimated and cross validated. The model predicted pain levels robustly and explained ∼36% of the pain variance. These results, afforded by fusing innovative analytical method with neuroimaging, shed light on the central mechanisms of TN and could inform future studies aimed at developing innovative therapies for treating TN.

## Data Availability

All data produced in the present study are available upon reasonable request to the authors.

## Acknowledgment

This study was supported by the Facial Pain Research Foundation.

## References

Ab Aziz, C.B., Ahmad, A.H., 2006. The role of the thalamus in modulating pain. The Malaysian journal of medical sciences: MJMS 13, 11.

Antonini, G., Di Pasquale, A., Cruccu, G., Truini, A., Morino, S., Saltelli, G., Romano, A., Trasimeni, G., Vanacore, N., Bozzao, A., 2014. Magnetic resonance imaging contribution for diagnosing symptomatic neurovascular contact in classical trigeminal neuralgia: a blinded case-control study and meta-analysis. PAIN® 155, 1464–1471.

Behrens, T.E., Johansen-Berg, H., Woolrich, M., Smith, S., Wheeler-Kingshott, C., Boulby, P., Barker, G., Sillery, E., Sheehan, K., Ciccarelli, O., 2003. Non-invasive mapping of connections between human thalamus and cortex using diffusion imaging. Nature neuroscience 6, 750–757.

Bennetto, L., Patel, N.K., Fuller, G., 2007. Trigeminal neuralgia and its management. Bmj 334, 201–205.

Bollimunta, A., Mo, J., Schroeder, C.E., Ding, M., 2011. Neuronal mechanisms and attentional modulation of corticothalamic alpha oscillations. Journal of Neuroscience 31, 4935–4943.

Borsook, D., Moulton, E.A., Pendse, G., Morris, S., Cole, S.H., Aiello-Lammens, M., Scrivani, S., Becerra, L.R., 2007. Comparison of evoked vs. spontaneous tics in a patient with trigeminal neuralgia (tic doloureux). Molecular pain 3, 1744–8069-1743-1734.

Bushnell, M.C., Čeko, M., Low, L.A., 2013. Cognitive and emotional control of pain and its disruption in chronic pain. Nature reviews neuroscience 14, 502–511.

Cacioppo, S., Frum, C., Asp, E., Weiss, R.M., Lewis, J.W., Cacioppo, J.T., 2013. A quantitative meta-analysis of functional imaging studies of social rejection. Scientific reports 3, 1–3.

Chen, Y., Xiang, C.Q., Liu, W.F., Jiang, N., Zhu, P.W., Ye, L., Li, B., Lin, Q., Min, Y.L., Su, T., 2019. Application of amplitude of low-frequency fluctuation to altered spontaneous neuronal activity in classical trigeminal neuralgia patients: A resting-state functional MRI study. Molecular medicine reports 20, 1707–1715.

Chen, Y., Yan, H., Han, Z., Bi, Y., Chen, H., Liu, J., Wu, M., Wang, Y., Zhang, Y., 2016. Functional activity and connectivity differences of five resting-state networks in patients with Alzheimer’s disease or mild cognitive impairment. Current Alzheimer Research 13, 234–242.

Coghill, R.C., Sang, C.N., Maisog, J.M., Iadarola, M.J., 1999. Pain intensity processing within the human brain: a bilateral, distributed mechanism. Journal of neurophysiology 82, 1934–1943.

Coppola, G., Di Renzo, A., Tinelli, E., Di Lorenzo, C., Scapeccia, M., Parisi, V., Serrao, M., Evangelista, M., Ambrosini, A., Colonnese, C., 2018. Resting state connectivity between default mode network and insula encodes acute migraine headache. Cephalalgia 38, 846–854.

Cunningham, S.I., Tomasi, D., Volkow, N.D., 2017. Structural and functional connectivity of the precuneus and thalamus to the default mode network. Human brain mapping 38, 938–956.

Davis, K.D., Taylor, S.J., Crawley, A.P., Wood, M.L., Mikulis, D.J., 1997. Functional MRI of pain-and attention-related activations in the human cingulate cortex. Journal of neurophysiology 77, 3370–3380.

De Ridder, D., Vanneste, S., 2016. Burst and tonic spinal cord stimulation: different and common brain mechanisms. Neuromodulation: Technology at the Neural Interface 19, 47–59.

DeCharms, R.C., Maeda, F., Glover, G.H., Ludlow, D., Pauly, J.M., Soneji, D., Gabrieli, J.D., Mackey, S.C., 2005. Control over brain activation and pain learned by using real-time functional MRI. Proceedings of the National Academy of Sciences 102, 18626–18631.

Ding, M., Bressler, S.L., Yang, W., Liang, H., 2000. Short-window spectral analysis of cortical event-related potentials by adaptive multivariate autoregressive modeling: data preprocessing, model validation, and variability assessment. Biological cybernetics 83, 35–45.

Ding, M., Chen, Y., Bressler, S.L., 2006. Granger causality: basic theory and application to neuroscience. Handbook of time series analysis: recent theoretical developments and applications, 437–460.

Dostrovsky, J.O., 2000. Role of thalamus in pain.

Echols, D.H., Colclough, J., 1947. Abolition of painful phantom foot by resection of the sensory cortex. Journal of the American Medical Association 134, 1476–1477.

Eisenberger, N.I., 2012. The pain of social disconnection: examining the shared neural underpinnings of physical and social pain. Nature reviews neuroscience 13, 421–434.

Fomberstein, K., Qadri, S., Ramani, R., 2013. Functional MRI and pain. Current Opinion in Anesthesiology 26, 588–593.

Freund, W., Klug, R., Weber, F., Stuber, G., Schmitz, B., Wunderlich, A.P., 2009. Perception and suppression of thermally induced pain: a fMRI study. Somatosensory & motor research 26, 1–10.

Freund, W., Stuber, G., Wunderlich, A.P., Schmitz, B., 2007. Cortical correlates of perception and suppression of electrically induced pain. Somatosensory & motor research 24, 203–212.

Frot, M., Mauguiere, F., Magnin, M., Garcia-Larrea, L., 2008. Parallel processing of nociceptive A-δ inputs in SII and midcingulate cortex in humans. Journal of Neuroscience 28, 944–952.

Granger, C.W., 1969. Investigating causal relations by econometric models and cross-spectral methods. Econometrica: journal of the Econometric Society, 424–438.

Greenspan, J.D., Lee, R.R., Lenz, F.A., 1999. Pain sensitivity alterations as a function of lesion location in the parasylvian cortex. Pain 81, 273–282.

Hagmann, P., Cammoun, L., Gigandet, X., Meuli, R., Honey, C.J., Wedeen, V.J., Sporns, O., 2008. Mapping the structural core of human cerebral cortex. PLoS biology 6, e159.

Huang, X., Zhang, D., Wang, P., Mao, C., Miao, Z., Liu, C., Xu, C., Yin, X., Wu, X., 2021. Altered amygdala effective connectivity in migraine without aura: evidence from resting‐state fMRI with Granger causality analysis. The journal of headache and pain 22, 1–8.

Ichesco, E., Puiu, T., Hampson, J., Kairys, A., Clauw, D., Harte, S., Peltier, S., Harris, R., Schmidt‐Wilcke, T., 2016. Altered fMRI resting‐state connectivity in individuals with fibromyalgia on acute pain stimulation. European journal of pain 20, 1079–1089.

Kim, H., 2010. Dissociating the roles of the default-mode, dorsal, and ventral networks in episodic memory retrieval. NeuroImage 50, 1648–1657.

Kim, J., Mawla, I., Kong, J., Lee, J., Gerber, J., Ortiz, A., Kim, H., Chan, S.-T., Loggia, M.L., Wasan, A.D., 2019. Somatotopically-specific primary somatosensory connectivity to salience and default mode networks encodes clinical pain. Pain 160, 1594.

Legrain, V., Iannetti, G.D., Plaghki, L., Mouraux, A., 2011. The pain matrix reloaded: a salience detection system for the body. Progress in neurobiology 93, 111–124.

Li, M., Yan, J., Li, S., Wang, T., Zhan, W., Wen, H., Ma, X., Zhang, Y., Tian, J., Jiang, G., 2017a. Reduced volume of gray matter in patients with trigeminal neuralgia. Brain imaging and behavior 11, 486–492.

Li, M., Yan, J., Li, S., Wang, T., Zhan, W., Wen, H., Ma, X., Zhang, Y., Tian, J., Jiang, G., 2017b. Reduced volume of gray matter in patients with trigeminal neuralgia. Brain imaging and behavior 11, 486–492.

Liang, Y., Zhao, Q., Hu, Z., Bo, K., Meyyappan, S., Neubert, J.K., Ding, M., 2022. Imaging the Neural Substrate of Trigeminal Neuralgia Pain Using Deep Learning. bioRxiv, 2022.2011.2002.514527.

Lieberman, M.D., Eisenberger, N.I., 2015. The dorsal anterior cingulate cortex is selective for pain: Results from large-scale reverse inference. Proceedings of the National Academy of Sciences 112, 15250–15255.

Loggia, M.L., Kim, J., Gollub, R.L., Vangel, M.G., Kirsch, I., Kong, J., Wasan, A.D., Napadow, V., 2013. Default mode network connectivity encodes clinical pain: an arterial spin labeling study. PAIN® 154, 24–33.

Maarbjerg, S., Wolfram, F., Gozalov, A., Olesen, J., Bendtsen, L., 2015. Significance of neurovascular contact in classical trigeminal neuralgia. Brain 138, 311–319.

Moisset, X., Villain, N., Ducreux, D., Serrie, A., Cunin, G., Valade, D., Calvino, B., Bouhassira, D., 2011. Functional brain imaging of trigeminal neuralgia. European journal of pain 15, 124–131.

Ning, Y., Zheng, R., Li, K., Zhang, Y., Lyu, D., Jia, H., Ren, Y., Zou, Y., 2018. The altered Granger causality connection among pain-related brain networks in migraine. Medicine 97.

Obermann, M., 2010. Treatment options in trigeminal neuralgia. Therapeutic advances in neurological disorders 3, 107–115.

Otti, A., Guendel, H., Läer, L., Wohlschlaeger, A.M., Lane, R.D., Decety, J., Zimmer, C., Henningsen, P., Noll-Hussong, M., 2010. I know the pain you feel—how the human brain’s default mode predicts our resonance to another’s suffering. Neuroscience 169, 143–148.

Porreca, F., Ossipov, M.H., Gebhart, G., 2002. Chronic pain and medullary descending facilitation. Trends in neurosciences 25, 319–325.

Porro, C.A., Cettolo, V., Francescato, M.P., Baraldi, P., 1998. Temporal and intensity coding of pain in human cortex. Journal of neurophysiology 80, 3312–3320.

Raichle, M.E., Gusnard, D.A., 2005. Intrinsic brain activity sets the stage for expression of motivated behavior. Journal of Comparative Neurology 493, 167–176.

Rainville, P., Duncan, G.H., Price, D.D., Carrier, B., Bushnell, M.C., 1997. Pain affect encoded in human anterior cingulate but not somatosensory cortex. science 277, 968–971.

Rajan, A., Meyyappan, S., Walker, H., Samuel, I.B.H., Hu, Z., Ding, M., 2019. Neural mechanisms of internal distraction suppression in visual attention. Cortex 117, 77–88.

Sestieri, C., Corbetta, M., Romani, G.L., Shulman, G.L., 2011. Episodic memory retrieval, parietal cortex, and the default mode network: functional and topographic analyses. Journal of Neuroscience 31, 4407–4420.

Seth, A.K., Barrett, A.B., Barnett, L., 2015. Granger causality analysis in neuroscience and neuroimaging. Journal of Neuroscience 35, 3293–3297.

Simons, L.E., Moulton, E.A., Linnman, C., Carpino, E., Becerra, L., Borsook, D., 2014. The human amygdala and pain: evidence from neuroimaging. Human brain mapping 35, 527–538.

Stramaglia, S., Angelini, L., Wu, G., Cortes, J.M., Faes, L., Marinazzo, D., 2016. Synergetic and redundant information flow detected by unnormalized Granger causality: Application to resting state fMRI. IEEE Transactions on Biomedical Engineering 63, 2518–2524.

Taylor, K.S., Seminowicz, D.A., Davis, K.D., 2009. Two systems of resting state connectivity between the insula and cingulate cortex. Human brain mapping 30, 2731–2745.

Tian, T., Guo, L., Xu, J., Zhang, S., Shi, J., Liu, C., Qin, Y., Zhu, W., 2016. Brain white matter plasticity and functional reorganization underlying the central pathogenesis of trigeminal neuralgia. Scientific reports 6, 1–11.

Trongnetrpunya, A., Nandi, B., Kang, D., Kocsis, B., Schroeder, C.E., Ding, M., 2016. Assessing granger causality in electrophysiological data: removing the adverse effects of common signals via bipolar derivations. Frontiers in systems neuroscience 9, 189.

Tsai, Y.H., Yuan, R., Patel, D., Chandrasekaran, S., Weng, H.H., Yang, J.T., Lin, C.P., Biswal, B.B., 2018. Altered structure and functional connection in patients with classical trigeminal neuralgia. Human brain mapping 39, 609–621.

Tu, Y., Jung, M., Gollub, R.L., Napadow, V., Gerber, J., Ortiz, A., Lang, C., Mawla, I., Shen, W., Chan, S.-T., 2019. Abnormal medial prefrontal cortex functional connectivity and its association with clinical symptoms in chronic low back pain. Pain 160, 1308.

Wakaizumi, K., Jabakhanji, R., Ihara, N., Kosugi, S., Terasawa, Y., Morisaki, H., Ogaki, M., Baliki, M.N., 2019. Altered functional connectivity associated with time discounting in chronic pain. Scientific reports 9, 1–11.

Wang, C., Rajagovindan, R., Han, S.-M., Ding, M., 2016a. Top-down control of visual alpha oscillations: sources of control signals and their mechanisms of action. Frontiers in human neuroscience 10, 15.

Wang, T., Chen, N., Zhan, W., Liu, J., Zhang, J., Liu, Q., Huang, H., He, L., Zhang, J., Gong, Q., 2016b. Altered effective connectivity of posterior thalamus in migraine with cutaneous allodynia: a resting-state fMRI study with granger causality analysis. The journal of headache and pain 17, 1–11.

Wang, Y., Zhang, X., Guan, Q., Wan, L., Yi, Y., Liu, C.-F., 2015. Altered regional homogeneity of spontaneous brain activity in idiopathic trigeminal neuralgia. Neuropsychiatric disease and treatment, 2659–2666.

Wen, X., Liu, Y., Yao, L., Ding, M., 2013a. Top-down regulation of default mode activity in spatial visual attention. Journal of Neuroscience 33, 6444–6453.

Wen, X., Rangarajan, G., Ding, M., 2013b. Is Granger causality a viable technique for analyzing fMRI data? PloS one 8, e67428.

Wen, X., Yao, L., Liu, Y., Ding, M., 2012. Causal interactions in attention networks predict behavioral performance. Journal of Neuroscience 32, 1284–1292.

Wiech, K., Jbabdi, S., Lin, C., Andersson, J., Tracey, I., 2014. Differential structural and resting state connectivity between insular subdivisions and other pain-related brain regions. PAIN® 155, 2047–2055.

Wilcox, C.E., Mayer, A.R., Teshiba, T.M., Ling, J., Smith, B.W., Wilcox, G.L., Mullins, P.G., 2015. The subjective experience of pain: an FMRI study of percept-related models and functional connectivity. Pain medicine 16, 2121–2133.

Wunderlich, A.P., Klug, R., Stuber, G., Landwehrmeyer, B., Weber, F., Freund, W., 2011. Caudate nucleus and insular activation during a pain suppression paradigm comparing thermal and electrical stimulation. The open neuroimaging journal 5, 1.

Xiang, C.Q., Liu, W.F., Xu, Q.H., Su, T., Yong‐Qiang, S., Min, Y.L., Yuan, Q., Zhu, P.W., Liu, K.C., Jiang, N., 2019. Altered spontaneous brain activity in patients with classical trigeminal neuralgia using regional homogeneity: a resting‐state functional MRI study. Pain Practice 19, 397–406.

Xu, H., Seminowicz, D.A., Krimmel, S.R., Zhang, M., Gao, L., Wang, Y., 2022. Altered structural and functional connectivity of salience network in patients with classic trigeminal neuralgia. The Journal of Pain 23, 1389–1399.

Yan, J., Li, M., Fu, S., Li, G., Wang, T., Yin, Y., Jiang, G., Lin, J., Li, W., Fang, J., 2019. Alterations of dynamic regional homogeneity in trigeminal neuralgia: a resting-state fMRI study. Frontiers in Neurology 10, 1083.

Yang, C.-X., Shi, T.-F., Liang, Q.-C., Yang, B.-F., Jiao, R.-S., Zhang, H., Zhang, Y., Xu, M.-Y., 2010. Cholecystokinin-8 antagonizes electroacupuncture analgesia through its B receptor in the caudate nucleus. Neuromodulation: Technology at the Neural Interface 13, 93–98.

Yin, Y., He, S., Xu, J., You, W., Li, Q., Long, J., Luo, L., Kemp, G.J., Sweeney, J.A., Li, F., 2020. The neuro-pathophysiology of temporomandibular disorders-related pain: a systematic review of structural and functional MRI studies. The journal of headache and pain 21, 1–20.

Zakrzewska, J.M., Linskey, M.E., 2014. Trigeminal neuralgia. Bmj 348.

Zhang, Z., Lu, G., Zhong, Y., Tan, Q., Liao, W., Wang, Z., Wang, Z., Li, K., Chen, H., Liu, Y., 2010. Altered spontaneous neuronal activity of the default-mode network in mesial temporal lobe epilepsy. Brain research 1323, 152–160.

Zhao, Z., Wang, X., Fan, M., Yin, D., Sun, L., Jia, J., Tang, C., Zheng, X., Jiang, Y., Wu, J., 2016. Altered effective connectivity of the primary motor cortex in stroke: a resting-state fMRI study with Granger causality analysis. PloS one 11, e0166210.

Zhu, P.-W., Chen, Y., Gong, Y.-X., Jiang, N., Liu, W.-F., Su, T., Ye, L., Min, Y.-L., Yuan, Q., He, L.-C., 2020. Altered brain network centrality in patients with trigeminal neuralgia: a resting-state fMRI study. Acta Radiologica 61, 67–75.

